# Occupational risk of SARS-CoV-2 infection and reinfection during the second pandemic surge: a cohort study

**DOI:** 10.1101/2021.08.06.21261419

**Authors:** Antonio Leidi, Amandine Berner, Roxane Dumont, Richard Dubos, Flora Koegler, Giovanni Piumatti, Nicolas Vuilleumier, Laurent Kaiser, Jean-François Balavoine, Didier Trono, Didier Pittet, François Chappuis, Omar Kherad, Delphine Courvoisier, Andrew S Azman, María-Eugenia Zaballa, Idris Guessous, Silvia Stringhini, SEROCoV-WORK+ study group

## Abstract

**Objectives:** This cohort study including essential workers, assessed the□risk and incidence of SARS-CoV-2□infection during the second surge of COVID-19 according to baseline serostatus and occupational sector.

**Methods:** Essential workers were selected from a seroprevalence survey cohort in Geneva, Switzerland and were linked to a state centralized registry compiling SARS-CoV-2 infections. Primary outcome was the number of virologically-confirmed infections from serological assessment (between May and September 2020) to January 25, 2021, according to baseline antibody status and stratified by three pre-defined occupational groups (occupations requiring sustained physical proximity, involving brief regular contact or others). Secondary outcomes included the incidence of infection.

**Results:** 10457 essential workers were included (occupations requiring sustained physical proximity accounted for 3057 individuals, those involving regular brief contact, 3645, and 3755 workers were classified under “Other essential occupations”). After a follow-up period of over 27 weeks, 5 (0.6%) seropositive and 830 (8.5%) seronegative individuals had a positive SARS-CoV-2 test, with an incidence rate of 0.2 (95% CI 0.1 to 0.6) and 3.2 (95% CI 2.9 to 3.4) cases per person-week, respectively. Incidences were similar across occupational groups. Seropositive essential workers had a 93% reduction in the hazard (HR of 0.07, 95% CI 0.03 to 0.17) of having a positive test during follow-up with no significant between-occupational group difference.

**Conclusions:** A ten-fold reduction in the hazard of being virologically tested positive was observed among anti-SARS-CoV-2 seropositive essential workers regardless of their sector of occupation, confirming the seroprotective effect of a previous SARS-CoV2 exposure at least six months after infection.

**Key messages:** *What is already known about this subject?:* Risk of SARS-CoV-2 reinfection is low in the general population and among healthcare workers.

*What are the new findings?:* A ten-fold reduction of risk of being virologically tested positive reinfection is observed among anti-SARS-CoV-2 seropositive essential workers of different activity sectors, regardless of their occupation-related risk of exposure.

*How might this impact on policy or clinical practice in the foreseeable future?:* Vaccination could be delayed in individuals with previous history of SARS-CoV-2 infection with serologic confirmation, regardless of their occupational exposure. These observations need to be confirmed for new SARS-CoV-2 variants.

## Introduction

The occupational risk of SARS-CoV-2 reinfection is incompletely understood. Large observational studies,^1 2^ especially among healthcare workers,^3^ found that natural infection elicits protective immunity with a ten-fold reduction of reinfection risk. Essential workers were unequally affected in the early phases of the pandemic, with healthcare workers (HCW) being at higher risk of contracting infections,^4 5^ and a wide variability in seropositivity across occupations.^6^ Close contact and inadequate personal protective equipment have been identified as risk factors,^7^ generating a large deployment of social distancing and barrier measures. Despite this, a second pandemic surge affected most countries worldwide, taking place at the end of 2020 in Switzerland. To face it, authorities in Geneva re-introduced partial lockdown policies, mandating home working and interrupting non-essential activities (Figure S1). As during the first lock down, these measures did not affect essential workers, whose occupations are considered indispensable for the society, such as workers in the healthcare, social work and transportation sectors. Workers in these sectors may face a higher risk of infection.^8^ During the first surge, HCW were at higher risk of infection but it remains unknown whether it is still the case with growing evidence that personal protective measures are effective,^9^ and if it also applies to the risk of reinfection. Little is known about the other essential occupations. We aimed therefore to investigate the protective effect conferred by previous infection, added and compared to the usual protective measures, in occupational groups with variable exposure risk. In this study, we assessed the risk of virologically-confirmed SARS-CoV-2 infection during the second pandemic surge in a large cohort of essential workers from 32 occupations according to their baseline serostatus and occupational group.

## Methods

Participants were selected from a serosurvey cohort recruiting essential workers between May and September 2020 in Geneva, Switzerland.^6^ Data of SARS-CoV-2 infections were extracted from a centralized state registry and linked to each participant, as previously described.^2^ Workers were categorised into three pre-defined groups, according to their exposure risk: occupations likely requiring sustained physical proximity to other individuals (e.g. HCW, childcare and social workers), occupations involving regular brief contact (e.g. pharmacists, taxi drivers, grocery workers) and other essential occupations (farmers, managers and health researchers) (Table S1). Participants were classified as seropositive or seronegative according to their serological status at recruitment (decisional algorithm available in supplementary material). SARS-CoV-2 infections were confirmed by reverse transcriptase polymerase chain reaction (RT-PCR) or antigenic rapid diagnostic test (Ag RDT) on naso-pharyngeal swabs.^2^ Positive RT-PCR or Ag RDT in seropositive individuals were clinically investigated by two independent adjudicators and classified as likely or unlikely reinfections. To note, no vaccine doses were available in Geneva during the study period.

### Outcomes

The primary outcome was the number of virologically-confirmed infections during study follow-up (i.e. from serological assessment to January 25, 2021) according to the baseline antibody status and stratified by occupational group. Secondary outcomes were the incidence of infections, incidence of testing and proportion of positive tests.

### Statistical analysis

The adjusted hazard ratio (HR) of having a virologically-confirmed infection in seropositive compared to seronegative participants was estimated with the Cox’s proportional hazard model. Variables having been previously associated to the risk of infection^10^ were included in the Cox’s model (age, sex, smoking status, obesity and formal educational level). Survival curves were presented using the Kaplan-Meier method and stratified by occupational group. Interaction between occupational group and serological status was tested with the likelihood ratio test. Consistency of results was assessed with the leave-one-out method where hazard ratios were computed by leaving each occupation out. Workers living outside the canton of Geneva, who may be tested at their living place, were excluded in a sensitivity analysis.

### Ethics

The investigation conforms the principles of the Declaration of Helsinki and was approved as an amendment by the local Ethical Committee (Commission Cantonale d’Éthique de la Recherche, Geneva, Switzerland; CCER 2020-00881). All participants gave written informed consent at the time of recruitment.

## Results

In total, 10582 essential workers from 32 occupations were included in the study cohort between May and September 2020 (57% and 55.6% of women and mean age of 43.9 and 44.5 years old for seropositive and seronegative individuals, respectively) (Tables S1 and S2). After exclusion of 125 participants with missing data in co-variables or outside the target age of 18 to 65 years old, occupations involving physical proximity accounted 3057 individuals, those involving regular customer contact 3645 individuals, and the others essential occupations 3755 individuals (Figure S2). Workers living in Geneva represented 57.5% of the study sample and were unequally distributed across occupational groups (Table S3). The follow-up period did not differ significantly between seropositive and seronegative participants, being 27.6 weeks (SD 5.2) and 27.9 (SD 5.1), respectively (*P*=0.061). On average, both seropositive and seronegative participants had 1.3 SARS-CoV-2 tests per individual during the study period with no differences between occupational groups (Table S3). Five (0.6%) seropositive and 830 (8.5%) seronegative individuals had a positive virologic SARS-CoV-2 test during the follow-up period. This corresponds to an incidence rate of 0.2 (95% CI 0.1 to 0.6) and 3.2 (95% CI 2.9 to 3.4) person-week for seropositive and seronegative individuals, respectively (Table S2). Incidences were similar across occupational groups (Table S3). All infections in seropositive individuals were considered likely reinfections by adjudicators (Table S4).

### Reinfection risk

Seropositive essential workers had a 93% reduction in the hazard of having a positive virologic SARS-CoV-2 test compared to those who were seronegative at baseline (HR of 0.07, 95% CI 0.03 to 0.17). No significant between-group difference was noted when stratifying by occupational group, the HR being 0.07 (95% CI 0.02 to 0.29) for occupations requiring physical proximity, 0.05 (95% CI 0.01 to 0.33) for occupations with regular customer contact, and 0.09 (95% CI 0.02 to 0.40) for other essential occupations (Figure 1 and Table S5, P_interaction_=0.85). Results were consistent in the leave-one-out sensitivity analysis (Figure S3) and in the subsample of participants living in the canton of Geneva (HR of 0.04, 95% CI 0.01 to 0.14, Table S6).

**Figure 1.**
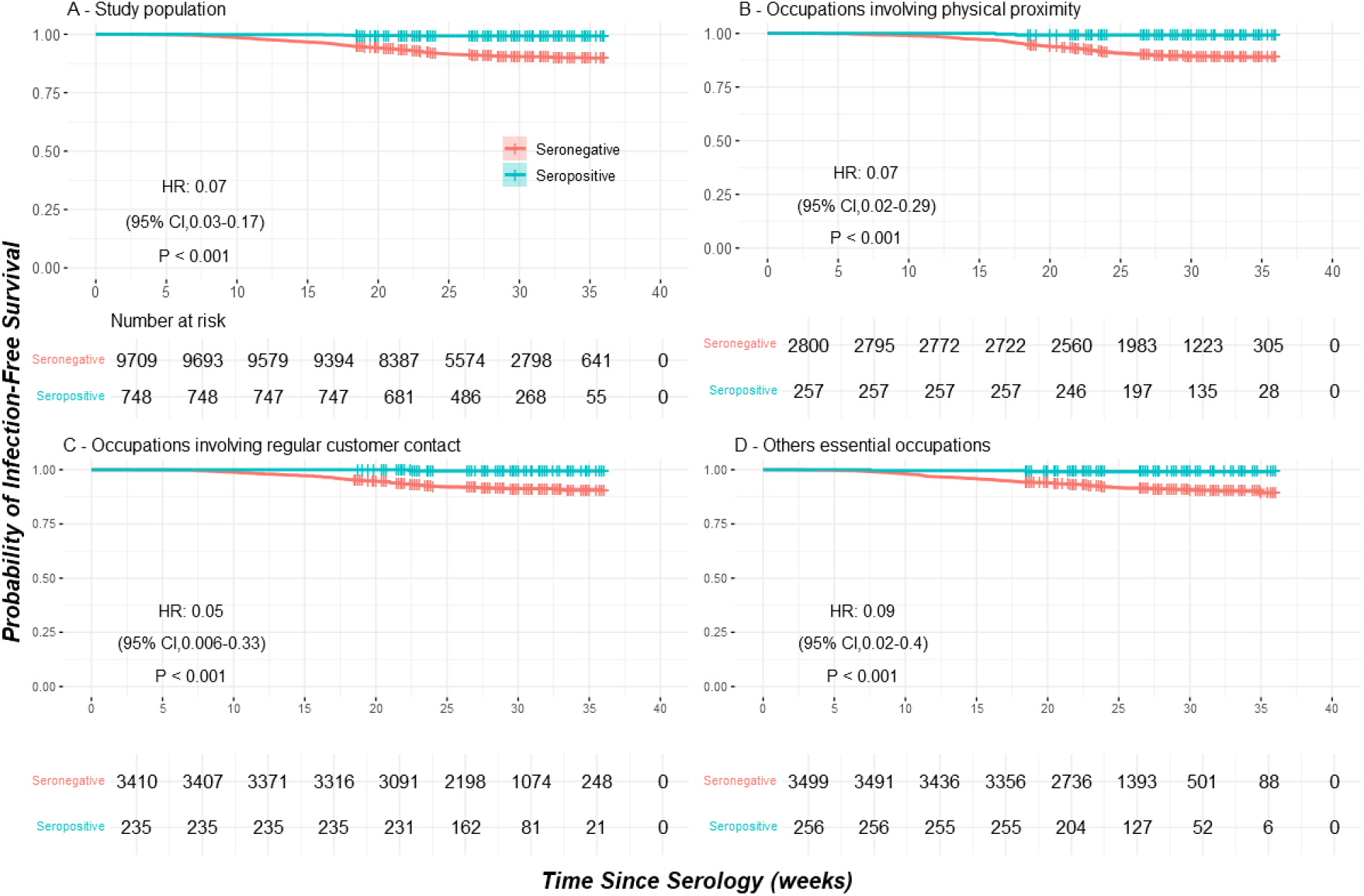
Infection-free survival according to serostatus in the study population (Panel A) and in subgroups of occupations (Panel B, C and D)

## Discussion

In this cohort study covering a period of over 27 weeks, a 93% reduction in the hazard of being virologically tested positive, was observed among anti-SARS-CoV-2 seropositive essential workers, compared to their seronegative counterparts, regardless of their sector of occupation and thus of the intensity of potential reexposure. This result is consistent with our report of a 94% risk reduction 35 weeks after serologic assessement in a previous cohort of the Geneva general population,^2^ as well as with a large retrospective analysis of RT-PCR results of the Danish population observing over 80% protection in the total study population and in the subgroup of HCW.^1^ Healthcare workers have been the focus of reinfection studies since they experienced higher rate of infections at the beginning of the pandemic. Based on these observations, HCW have been prioritised by vaccination policies, even if already been infected. Other occupations however, such as waiters and taxi drivers, were possibly at higher risk for infection than HCW during the second phase of pandemic.^11^ The present study observed a similar incidence of infection in seronegative individuals across occupational groups with variable exposure risk, suggesting the effectiveness of universal implementation of personal protection policies. Moreover, we observed a similar reinfection rate across the occupational groups, suggesting protection against reinfection, regardless of the various degree of exposure.

The study has some limitations. First, essential workers were categorized into three pre-specified groups, based on previous reports on exposure risk, though these groupings are imperfect and may not entirely reflect exposure.^4^ Second, our sample was composed by a significant proportion of cross-border workers, possibly being tested outside Geneva. Similar results, however, were observed when border workers were excluded (HR of 0.04, 95%CI 0.01 to 0.14). Third, participants of the SEROCoV-WORK+ were included on a voluntary basis, and selection towards participants better aware of sanitary measures might have occurred, potentially underestimating reinfection risk and limiting generalizability of results. Finally, we cannot infer cross-protection against new SARS-CoV-2 variants because they were underrepresented during the study period.

This study has several important strengths. First, serological status assessment occurred in the early phases of the pandemic allowing a longitudinal follow-up covering the second pandemic surge. Second, this study took place before vaccination era, consenting us to properly evaluate the protective effect conferred by natural infection. Third, a large number of essential occupations were represented, providing information outside the healthcare sector. Finally, study results were concordant in all sensitivity analyses raising robustness of our observations.

In conclusion, we observed no significant differences in documented SARS-CoV-2 infections and reinfections across mobilized workers from a variety of occupational categories. Our results suggest the efficiency of universal barrier measures deployment and build up on the growing evidence about the seroprotective effect of antibodies at least six months after infection.

## Data Availability

All data is available for consultation

## Acknowledgments

We would like to thank Aglaé Tardin and the team of General Directorate of Health of Geneva for giving us access to the ARGOS state registry; We thank the Hôpital de la Tour, the Hirslanden Clinique des Grangettes, and the Hirslanden Clinique de la Colline for participating as testing centers and the Geneva Chamber of Commerce, Industry, and Services (CCIG) for contributing to the recruitment of facilities for SEROCoV-WORK+ study. We thank all participants, whose contributions were invaluable and integral to the study.

## Author contributorship

The idea for the study originally came from AL. AL and AB carried out the literature search. AL, AB, SS, IG, RDum, RDub, ASA and MEZ conceptualized and designed the study. GP and DC oversaw database linkage. AB and FK conducted clinical investigations. RDum, RDub did data analysis. AL, AB, SS, ASA wrote the first draft of the manuscript. All authors have read, critically revised and approved the final version of this manuscript. AL and AB, SS and IG contributed equally to this paper.

## Competing Interests

None to declare.

## Funding

This study was funded by the Private Foundation of the Geneva University Hospitals, the Fondation des Grangettes and the Center for Emerging Viral Diseases.

## Notes

### Competing Interest Statement

The authors have declared no competing interest.

### Author Declarations

The investigation conforms the principles of the Declaration of Helsinki and was approved as an amendment by the local Ethical Committee (Commission Cantonale d'Ethique de la Recherche, Geneva, Switzerland; CCER 2020-00881). All participants gave written informed consent at the time of recruitment.

