## Supplementary material for "Occupational risk of SARS-CoV-2 infection and reinfection during the second pandemic surge: a cohort study": Online Supplementary Materials

Antonio Leidi^1*^ MD, Amandine Berner^1*^ MD, Roxane Dumont^2^, Richard Dubos^2^, Flora Koegler^1^ MD, Giovanni Piumatti^2,3^ PhD, Prof Nicolas Vuilleumier^4^ MD, Prof Laurent Kaiser^5^ MD, Prof Jean-François Balavoine^6^, Prof Didier Trono^7^, Prof Didier Pittet^8^, Prof François Chappuis^2^, Prof Omar Kherad^9^, Prof Delphine Courvoisier^10^ PhD, Andrew S Azman^2.11^ PhD, María-Eugenia Zaballa^2^ PhD, Prof Idris Guessous^2*^ MD, Prof Silvia Stringhini^2*^ PhD, SEROCoV-WORK+ study group

*These authors contributed equally to the study

^1^Division of General Internal Medicine, Geneva University Hospitals, Geneva, Switzerland

^2^Division of Primary Care Medicine, Geneva University Hospitals, Geneva, Switzerland

^3^Institute of Public Health, Faculty of BioMedical Sciences, Università della

Svizzera Italiana, Lugano, Switzerland

^4^Division of Laboratory Medicine, Geneva University Hospitals, Geneva, Switzerland

^5^Geneva Center for Emerging Viral Diseases, Geneva University Hospitals, Geneva, Switzerland

^6^Department of Medicine, Faculty of Medicine, University of Geneva, Switzerland

^7^School of Life Sciences, Ecole Polytechnique Fédérale de Lausanne (EPFL), Lausanne, Switzerland

^8^Infection Control Program and World Health Organization Collaborating Center on Patient Safety, Geneva University Hospitals and Faculty of Medicine, Geneva, Switzerland.

^9^Division of Internal Medicine, Hôpital de la Tour and Faculty of Medicine, Geneva, Switzerland

^10^General Directorate of Health, Geneva, Switzerland

^11^Department of Epidemiology, Johns Hopkins Bloomberg School of Public Health, Baltimore, USA.

**Members of the SEROCoV-WORK + Study Group**

Victoria Alber, Isabelle Arm-Vernez, Andrew S. Azman, Delphine Bachmann, Donatien Bachmann, Stéphanie Baggio, Jean-François Balavoine, Gil Barbosa Monteiro, Hélène Baysson, Patrick Bleich, Isabelle Boissel, François Chappuis, Prune Collombet, Delphine Courvoisier, Philippine Couson, Alioucha Davidovic, Clement Deiri, Divina Del Rio, Carlos de Mestral, David De Ridder, Yaron Dibner, Paola D’Ippolito, Joséphine Duc, Roxane Dumont, Isabella Eckerle, Nacira El Merjani, Gwennaelle Ferniot, Antonie Flahault, Natalie Francioli, Marion Frangville, Carine Garande, Laurent Gétaz, Pamela Giraldo, Fanny Golaz, Julie Guérin, Idris Guessous, Ludivine Haboury, Séverine Harnal, Victoria Javet, Laurent Kaiser, Omar Kherad, Amélie Laboulais, Gaëlle Lamour, Xavier Lefebvre, Pierre Lescuyer, Andrea Jutta Loizeau, Fanny-Blanche Lombard, Elsa Lorthe, Chantal Martinez, Kourosh Massiha, Ludovic Metral-Boffod, Benjamin Meyer, Khaled Mostaguir, Mayssam Nehme, Natacha Noël, Nicolas Oederlin, Francesco Pennacchio, Javier Perez-Saez, Dusan Petrovic, Attilio Picazio, Didier Pittet, Giovanni Piumatti, Jane Portier, Géraldine Poulain, Caroline Pugin, Nick Pullen, Barinjaka Rakotomiaramanana, Zo Francia Randrianandrasana, Aude Richard, Viviane Richard, Sabina Rodriguez-Velazquez, Lilas Salzmann-Bellard, Silvia Stringhini, Leonard Thorens, Simon Torroni, Didier Trono, David Vidonne, Guillemette Violot, Nicolas Vuilleumier, Zoé Waldmann, Manon Will, Ania Wisniak, Sabine Yerly & María-Eugenia Zaballa

**Sequential serology assessment algorithm**

Seropositivity was first assessed by the detection of IgG antibodies against the S1 domain of SARS-CoV-2 spike protein using a commercially available ELISA (Euroimmun, Lübeck, Germany, #EI 2606-9601 G, cut-off for positivity ≥4.0, for negativity <0.8). Cases with intermediate results were tested for total Ig antibodies (IgG/A/M) against the virus nucleocapside protein using the Elecsys® anti-N assay (Roche Diagnostics, Rotkreuz, Switzerland, #09 203 079 190, cut-off for positivity >1.1, for negativity <0.8). Finally, still indeterminate cases were subject to a recombinant immunofluorescence assay (rIFA). This algorithm was designed to minimize false positive and false negative results at the individual level based on commercially available tests at the time of recruitment as well as practical constrains given the large number of participants.

**Figure S1. Weekly incidence of virologically-confirmed SARS-CoV-2 infections in the canton of Geneva, Switzerland, between March 2020 and February 2021.**

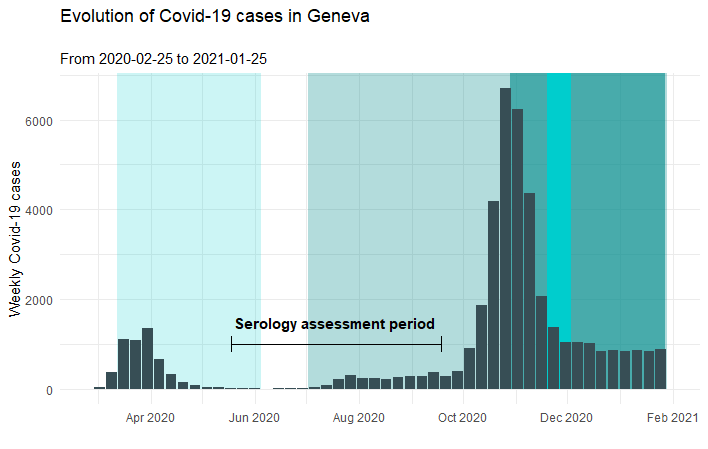

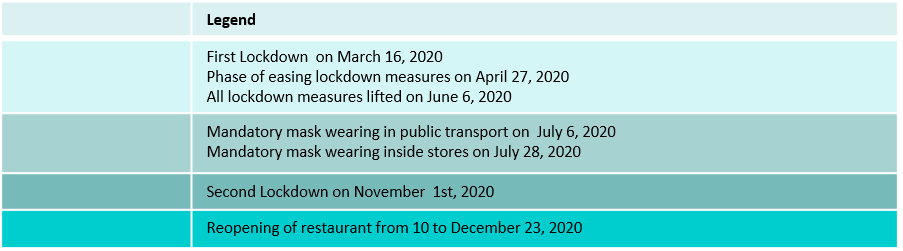

**Table S1. Description of occupations**

| Occupation | N | Description |
| --- | --- | --- |
| Occupations involving sustained physical proximity | **3057** |  |
| Doctor/surgeon | 369 | Medical doctor and surgeon |
| Nurse/assistant nurse | 1521 | Nurse and assistant nurse |
| Firefighter/emt | 275 | Firefighter, first aid responder, EMT |
| Domestic care worker | 129 | In-home caregiver, personal care assistant, companion, housekeeper |
| Childcare worker | 185 | Childcare worker, childcare educator, assistant childcare educator |
| Social worker | 578 | Social worker, assistant social worker, family worker, youth worker, etc. |
| Occupations involving regular brief contact | **3645** |  |
| Pharmacist/assistant pharmacist | 206 | Pharmacist, assistant pharmacist, pharmacy apprentice/intern |
| Other healthcare personnel | 849 | Other healthcare professional, biomedical/laboratory technician |
| Cashier | 138 | Cashier /sales worker |
| Public transport driver | 345 | Bus/tram driver, reduced mobility person driver, ticket controller |
| Taxi driver/chauffeur | 36 | Taxi driver, chauffeur |
| Gas station worker | 14 | Gas station worker/personnel |
| Police officer | 696 | Police officer or gendarme, civil service, army/soldier, civil protection |
| Bank/post office teller | 100 | Teller/counter agent in banks and post offices |
| Banking reception | 99 | Receptionist at banks |
| Teacher | 35 | Teacher, teaching assistant, tutor |
| Supermarket/food market personnel | 255 | Personnel working in supermarkets or small food markets/stores |
| Construction and food craft/trades | 496 | Construction worker, mechanic, metal worker, artisan, watch maker, electrician, electro-mechanic, warehouse worker, handler, repair person; food-processing worker, butcher, slaughterhouse worker, dairy worker, cheesemaker; hairdresser |
| Undertaker | 39 | Undertaker, funeral home agent, gravedigger |
| Kitchen staff | 159 | Cook, assistant cook, kitchen helper, food server, etc. |
| Journalist | 178 | Journalist, photographer, camera person, stage manager, etc. |
| Other essential occupations | **3755** |  |
| Home delivery driver | 42 | Delivery driver, courier, dispatch rider |
| Manager/assistant manager | 418 | Manager and assistant manager in customer service roles, apprentice/intern in customer service |
| Cleaner | 325 | Cleaning, washing, and maintenance service worker |
| Security guard | 205 | Security or surveillance agent, night watch, etc. |
| Lorry driver | 32 | Truck/lorry long-haul driver |
| Farmer/gardener | 104 | Farmer, nursery owner, gardener, horticulturalist, winegrower, winemaker |
| Health researcher/research personnel | 27 | Healthcare researcher, research associate, research assistant |
| Administration/hr | 1606 | Administrative, human resources, secretary, accounting employee |
| Communication/marketing manager | 104 | Communication or marketing manager, assistant manager |
| Finance, management, law, engineering | 733 | Engineer, data analysts, statistician, computer engineer/specialist, IT support; financial analyst, business analyst, cash management technical officer, compliance officer, accountant, consultant, fiscal controller, architect, civil engineer, investor; lawyer, court clerk, legal expert, judge, magistrate, prosecutor, air traffic controller, flight coordinator, chemist |
| Other professions | 159 |  |

|  | **Seropositive (n=748)** | **Seronegative (n=9709)** | ***P* Value** |
| --- | --- | --- | --- |
| Age, years, mean (SD) | 43.9 (10.9) | 44.5 (10.6) | 0.185 |
| Female, No. (%) | 423 (57.0) | 5399 (55.6) | 0.664 |
| BMI, kg/m^2^, mean (SD) | 25 (4.2) | 25 (4.4) | 0.956 |
| Education, No. (%)  Lower education  Middle education  Higher education | 69 (9)  297 (40)  382 (51) | 784 (8)  4285 (44)  4640 (47) | 0.055 |
| Smoking status, No. (%)  Non-smoker  Ex-smoker  Smoker | 519 (70)  109 (15)  120 (15) | 5573 (58)  1551 (16)  2585 (26) | <0.001 |
| Follow-up, weeks, mean (SD) | 27.6 (5.2) | 27.9 (5.1) | 0.061 |
| Incidence of testing after study inclusion, No. per individual, mean (SD) | 1.3 (0.6) | 1.3 (0.7) | 0.153 |
| Proportion of positive test during follow-up, % | 2.2% | 19.0% | <0.001 |
| Incidence of virologically confirmed SARS-CoV-2 infections, No. every 1000 person-week (95%CI) | 0.2 (0.1 to 0.6) | 3.2 (2.9 to 3.4) | <0.001 |

**Table S2. Characteristics of the study individuals according to baseline antibody status**

|  | Occupations involving sustained physical proximity (n= 3057) | | Occupations involving regular brief contact (n= 3645) | | Other essential occupations (n=3755) | |
| --- | --- | --- | --- | --- | --- | --- |
|  | **Seropositive (n= 257)** | **Seronegative (n=** **2800)** | **Seropositive (n= 235)** | **Seronegative (n= 3410)** | **Seropositive (n=** **256)** | **Seronegative (n=** **3499)** |
| Age, years, mean (SD) | 42.2 (11.3) | 43.4 (10.7) | 42.4 (10.9) | 43.7 (11) | 47 (9.7) | 46.2 (10.1) |
| Female, No. (%) | 197 (77) | 2084 (74.0) | 100 (42.6) | 1503 (44) | 126 (49.2) | 1812 (51.8) |
| BMI group, No. (%)  < 25 kg/m^2^  25-29.9 kg/m^2^  ≥ 30 kg/m^2^ | 157 (61)  70 (27)  30 (12) | 1788 (63)  743 (27)  269 (10) | 129 (55)  79 (33)  27 (12) | 1819 (53)  1152 (34)  439 (13) | 149 (58)  79 (30)  28 (12) | 1934 (56)  1102 (31)  463 (13) |
| Education, No. (%)  Lower education  Middle education  Higher education | 15 (6)  85 (33)  157 (61) | 175 (6)  987 (35)  1638 (59) | 34 (15.0)  122 (52.0)  79 (33.0) | 342 (10.0)  1915 (56.0)  1153 (34.0) | 20 (7.8)  90 (35.2)  146 (57) | 267 (8)  1383 (40)  1849 (54) |
| Smoking status, No. (%)  Non-smoker  Ex-smoker  Smoker | 185 (72)  31 (12)  41 (16) | 1569 (56)  447 (16)  784 (28) | 167 (71)  36 (15)  32 (14) | 1916 (56)  541 (16)  953 (28) | 167 (66)  42 (15)  47 (18) | 2088(65)  563 (16)  848 (19) |
| Resident in Geneva, No. (%) | 124 (48.2) | 1381 (49.3) | 131 (55.7) | 1902 (55.8) | 169 (66.0) | 2268 (64.8) |
| Chronic condition, No. (%) | 38 (15) | 311 (11.1) | 35 (15) | 413 (12) | 34 (13) | 447 (13) |
| Follow-up, weeks, mean (SD) | 29.6 (5) | 28.4 (6) | 28.4 (4.7) | 27.4 (5.8) | 25.7 (5.2) | 24.4 (5.7) |
| Rate of testing after study inclusion, No. per individual, mean (SD) | 1.3 (0.8) | 1.4 (0.8) | 1.2 (0.5) | 1.3 (0.6) | 1.3 (0.6) | 1.3 (0.7) |
| Proportion of positive test during follow-up, % | 1.9% | 17.5% | 1.9% | 21% | 2.8% | 19% |
| Incidence of virologically confirmed SARS-CoV-2 infections, No. every 1000 person-week (95%CI) | 0.26 (0.03 to 0.95) | 3.5 (3.1 to 3.9) | 0.14 (0.2 to 0.8) | 2.96 (2.6 to 3.3) | 0.3 (0.04 to 1.1) | 3.19 (2.8 to 3.6) |

**Table S3. Characteristics of the study individuals according to antibody status and occupational category**

**Figure S2. Study flow diagram**. Positive test refers to COVID-19 virological diagnostic test during follow-up**.**

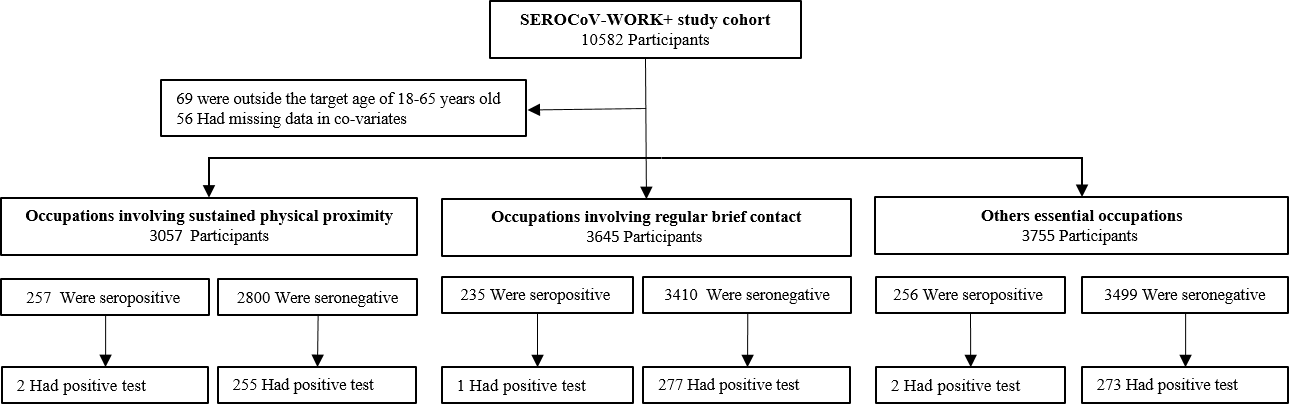

**Table S4. Demographic, clinical and laboratory characteristics of reinfected seropositive individuals**

| Patients | | Baseline serologic assay  (EI, IgG ratio) | No of days between serologic assay and potential reinfection | Clinical characteristics | Timing of PCR, CT value | Reinfection^a^ | |
| --- | --- | --- | --- | --- | --- | --- | --- |
|  |  |  |  |  |  | OA1 | OA2 |
| 1 | **female**  **between 36 and 40 years old (y.o.)** | positif  8.29 | 53 | 1st episode: asymptomatic    2nd episode: symptomatic (mild COVID-19 - like symptoms: fever, sore throat, anosmia, dysgeusia, headache, diarrhea) | 1st episode: not done (asymptomatic)    2nd episode: PCR (Cobas 6800 assay), CT value 23.8 | likely | likely |
| 2 | **Female**  **between 31-35 y.o.** | positif  6.82 | 113 | No information | 1st episode: no information    2nd episode: PCR, no CT value | likely | likely |
| 3 | **female**  **Between 26 and 30 y.o.** | positif  4.41 | 123 | No information | 1st episode: no information    2nd episode: PCR, no CT value | likely | likely |
| 4 | **male**  **between 46 and 50 y.o.** | positif  6.71 | 129 | 1st episode: asymptomatic    2nd episode: symptomatic (mild COVID-19 - like symptoms: fever, chills, headache, fatigue) | 1st episode:  not done (asymptomatic)    2nd episode: PCR (Cobas 6800 assay), CT value 27.1 | likely | likely |
| 5 | **Female**  **between 46 and 50 y.o.** | positif  6.43 | 157 | 1st episode: cough, fever, fatigue by the end of december 2019^b^. No history of recent travel outside Switzerland, no proven contact with a covid-19 case.    2nd episode: symptomatic (mild COVID-19 - like symptoms: fever, chills, myalgia & arthralgia, headache, fatigue) | 1st episode: not done    2nd episode: PCR, no CT value | likely | likely |

^a^Two adjudicators evaluated independently the reinfection probability of each case.

^b^In Switzerland, the first COVID-19 case was reported on February 25th, 2020.

EI means Euroimmun anti-S1 SARS-CoV-2 IgG ELISA. PCR means Polymerase Chain Reaction. CT means Cycle Threshold. OA means Outcome Adjudicator.

**Table S5. Number and risk of infection in seropositive and seronegative participants for the total study population and occupational categories.**

| Groups | **Number of infections, No. (%)** | | **Hazard ratio (95%CI)** | ***P* value** |
| --- | --- | --- | --- | --- |
|  | **seropositive** | **seronegative** |  |  |
| Study population (n= 10457) | 5 (0.6) | 830 (8.5) | 0.07 (0.03 to 0.17) | <0.001 |
| Occupations involving sustained physical proximity (n=3057) | 2 (0.8) | 280 (9.1) | 0.07 (0.02 to 0.29) | <0.001 |
| Occupations involving regular brief contact (n= 3645) | 1 (0.4) | 277 (8) | 0.05 (0.01 to 0.33) | <0.001 |
| Others essential occupations (n=3755) | 2 (0.8) | 273 (7.8) | 0.09 (0.02 to 0.40) | <0.001 |

**Figure S3. Leaving-one-profession-out estimation of hazard ratio infection in seropositive compared to seronegative essential workers**

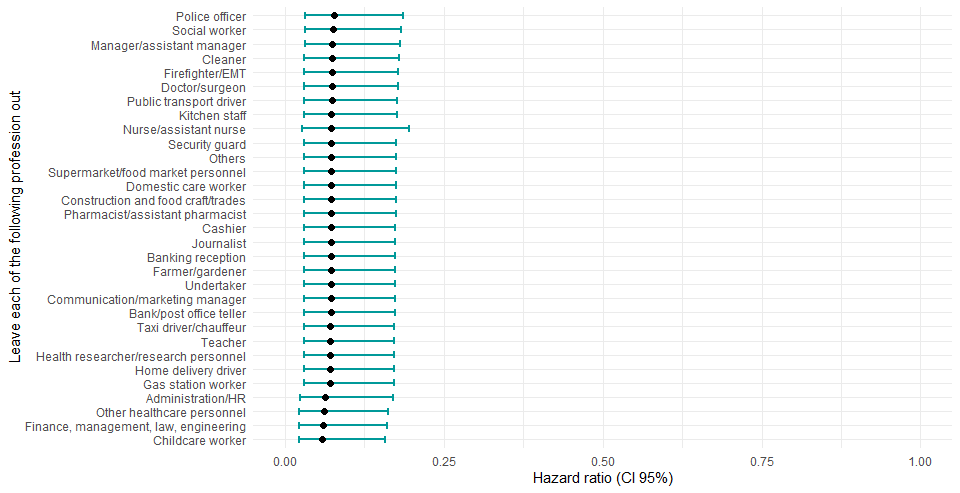

**Table S6. Number and risk of infection in seropositive and seronegative participants of the total study population and occupational category subgroups among participants living in Geneva.**

| Groups | **Number of infections, No. (%)** | | **Hazard ratio (95%CI)** | ***P* value** |
| --- | --- | --- | --- | --- |
|  | **seropositive** | **seronegative** |  |  |
| Geneva residents (n= 5975) | 2 (0.47) | 632 (11.3) | 0.04 (0.01, 0.14) | <0.001 |
| Occupations involving sustained physical proximity (n=1505) | 0 | 181 (13) | 0 (0) undefined | <0.001 |
| Occupations involving regular brief contact (n=2033) | 0 | 216 (11.3) | 0 (0) undefined | <0.001 |
| Others essential occupations (n=2437) | 2 (1.2) | 235 (10.3) | 0.1 (0.03, 0.4) | <0.001 |
